# Non-pharmacological therapies for post-viral syndromes, including Long COVID: A systematic review

**DOI:** 10.1101/2022.06.07.22276080

**Authors:** Joht Singh Chandan, Kirsty R Brown, Nikita Simms-Williams, Nasir Z Bashir, Jenny Camaradou, Dominic Heining, Grace M Turner, Samantha Cruz Rivera, Richard Hotham, Sonica Minhas, Krishnarajah Niratharakumar, Manoj Sivan, Kamlesh Khunti, Devan Raindi, Steven Marwaha, Sarah E. Hughes, Christel McMullan, Tom Marshall, Melanie J Calvert, Shamil Haroon, Olalekan Lee Aiyegbusi, the TLC Study Group

**Author notes:** **Corresponding author:** Dr Joht Singh Chandan, Institute of Applied Health Research, University of Birmingham, Birmingham, UK. Joint senior author (Shamil Haroon, Olalekan Lee Aiyegbusi). **Author statement:** The research questions were conceived by JSC, KN, MJC and SH. JSC, KB, NSM, DH, OLA, GT, SCR, RH, SEH, CM and SH all contributed to screening and data extraction steps. JSC and KB contributed to the initial draft of the manuscript. Further reviews and revisions to the manuscript were made by all remaining co-authors (JC, SM, NB, KN, MS, KK, DR, TM, SM and MJC). The final copy of the manuscript was approved by all authors. **Data sharing:** The data used as part of this review can be requested from the corresponding author. **Ethics approval:** This study does not require ethical approval as it is a secondary analysis of published data. **Transparency statement:** The lead author (the manuscript’s guarantor) affirms that this manuscript is an honest, accurate, and transparent account of the study being reported; that no important aspects of the study have been omitted; and that any discrepancies from the study as planned (and, if relevant, registered) have been explained.

## Abstract

**Background:** Post-viral syndromes (PVS), including Long COVID, are symptoms sustained from weeks to years following an acute viral infection. Non-pharmacological treatments for these symptoms are poorly understood. This review summarises evidence for the effectiveness of non-pharmacological treatments for symptoms of PVS. It also summarises the symptoms and health impacts of PVS in individuals recruited to studies evaluating treatments.

**Methods and findings:** We conducted a systematic review to evaluate the effectiveness of non-pharmacological interventions for PVS, as compared to either standard care, alternative non-pharmacological therapy, or placebo. The outcomes of interest were changes in symptoms, exercise capacity, quality of life (including mental health and wellbeing), and work capability. We searched five databases (Embase, MEDLINE, PsycINFO, CINAHL, MedRxiv) for randomised controlled trials (RCTs) published between 1^st^ January 2001 to 29^th^ October 2021. We anticipated that there would be few RCTs specifically pertaining to Long COVID, so we also included observational studies only if they assessed interventions in individuals where the viral pathogen was SARS-COV-2. Relevant outcome data were extracted, study quality appraised using the Cochrane Risk of Bias tool, and the findings were synthesised narratively. Quantitative synthesis was not planned due to substantial heterogeneity between the studies. Overall, five studies of five different interventions (Pilates, music therapy, telerehabilitation, resistance exercise, neuromodulation) met the inclusion criteria. Aside from music-based intervention, all other selected interventions demonstrated some support in the management of PVS in some patients.

**Conclusions:** In this study, we observed a lack of robust evidence evaluating non-pharmacological treatments for PVS, including Long COVID. Considering the prevalence of prolonged symptoms following acute viral infections, there is an urgent need for clinical trials evaluating the effectiveness and cost-effectiveness of non-pharmacological treatments for patients with PVS as well as what may work for certain sub-groups of patients with differential symptom presentation.

**Registration:** The study protocol was registered with PROSPERO [CRD42021282074] in October 2021 and published in BMJ Open in 2022.

**Author summary:** Why was this study done?

- The prevalence of Long COVID following exposure to SARS CoV-2 is substantial, and the current guidance provides few evidence-based treatment options for clinicians to suggest to their patients.
- Due to the similarities in presentation of other post-viral syndromes (PVS), and the lack of consensus in management approaches, there is a need to synthesise the available data on PVS to both support patients with PVS predating the pandemic, and those with Long COVID.

What did the researchers do and find?

- This is the first comprehensive systematic review of the effectiveness of non-pharmacological treatments for patients with PVS, including Long COVID.
- We identified four non-pharmacological treatments (Pilates, telerehabilitation, resistance exercises and neuromodulation) which have shown promise in those who have experienced signs and symptoms related to PVS.

What do these findings mean?

- In this study, we identified few trials assessing the effectiveness of non-pharmacological therapies to support the management of symptoms of PVS. Considering the prevalence of PVS, including Long COVID, there is an urgent need for clinical trials evaluating the effectiveness and cost-effectiveness of non-pharmacological therapies to support these patients.

## Background

Globally, there have been over 520 million cases of COVID-19, with over 6 million associated deaths, as of 13 May 2022.^1^ The pandemic has triggered a concerted global effort to rapidly develop and deliver safe vaccinations at record speed, which have significantly reduced morbidity, mortality, and disease transmission associated with Severe Acute Respiratory Syndrome Coronavirus-2 (SARS CoV-2) infection.^2–4^ Although vaccines have substantially reduced mortality, the breadth and relapsing-remitting nature of ongoing symptoms (such as fatigue and dyspnoea) that may arise from even mild infections of COVID-19 pose a substantial burden to patients and health services.^5,6^ The term ‘Long COVID’ used throughout this review is the generally preferred terminology for such ongoing symptoms following COVID-19 infection, known also as post-COVID-19 condition or post-acute sequelae of SARS-CoV-2 infection (PASC).^7,8^

According to ONS data, 1.7 million people living in private households in the United Kingdom (UK) (2.7% of the population) report a self-diagnosis of Long COVID, with 1.1 million (71%) having confirmed or suspected COVID-19 more than 12 weeks prior.^5^ Despite the substantial prevalence of these ongoing symptoms, the current National Institute for Health and Care Excellence (NICE) guideline for the management of the Long COVID provides only limited guidance for patients, focusing on self-management, individual goal setting through shared decision making, phased returns to work for those capable and rehabilitation.^9^

Although, the nature and presentations of Long COVID is still under investigation, many of its clinical features, including respiratory, neurological, psychological and gastroenterological complications, and specifically fatigue, occur following other acute viral infections.^10,11^ Such prolonged sequelae are referred to as post-viral syndromes (PVS) and follow exposure to viral pathogens, such as Epstein-Barr Virus (EBV) and Chikungunya Virus.^12^ The effectiveness of therapies for prolonged symptoms, such as fatigue, following infection with endemic viruses like EBV, have previously been investigated.^13,14^

A recent mixed-methods systematic review of post-viral fatigue syndromes provides useful lessons that may guide the development of Long COVID support services.^14^ However, to our knowledge, beyond the synthesis of interventions used to support patients with fatigue, there is no review of the wider array of non-pharmacological interventions for patients with symptoms such as dyspnoea or arthralgia, both common with Long COVID.^6^ Therefore, there is an urgent need to identify whether the existing evidence base on similar PVS can inform the management of patients with Long COVID. Hence, this systematic review summarises the evidence on non-pharmacological treatments for PVS, including Long COVID.

## Methods

This review has been conducted in accordance with the PRISMA guidelines (Supplementary table 1).^15^ The protocol was registered on PROSPERO (CRD42021282074) in October 2021 and the full protocol has been published elsewhere.^16^

### Eligibility criteria

#### Population

The population of interest consisted of adults and children with a PVS (including Long COVID). As there is no agreed definition of PVS, there is heterogeneity in the temporal description of PVS onset following the initial viral exposure. In line with the minimum timeframe for Long COVID used by the World Health Organization (WHO), where temporal criteria were described, we included only those studies where post-viral symptoms lasted beyond 12 weeks.^17^ However, we also included publications that provided no firm timeframe but indicated an aspect of chronicity or prolonged persistence of symptoms.

#### Intervention and comparator

We included studies that assessed the effectiveness of non-pharmacological interventions designed to improve symptoms of PVS against standard care, an alternative non-pharmacological therapy, or a placebo.

#### Outcomes

The outcomes were changes in symptoms, exercise capacity, quality of life (including changes in mental and physical wellbeing) and work capability.

#### Study type

Only randomised controlled trials (RCT) were included for patients with PVS, for conditions other than COVID-19. However, as we anticipated lack of RCTs for SARS-CoV-2, we also included observational studies where the viral pathogen was SARS-CoV-2.

We included primary research studies published between 1^st^ January 2001 and 29th October 2021. These dates were chosen to encompass research on the first Severe Acute Respiratory Syndrome (SARS) and Middle East Respiratory Syndrome (MERS) outbreaks which are the two pandemic viruses most related to SARS-CoV-2. There were no restrictions on setting (i.e., community or hospital based) or language.

### Information sources and search strategy

Four electronic databases were initially searched for RCTs evaluating interventions for PVS: Embase, MEDLINE, PsycINFO, and Cumulative Index to Nursing and Allied Health Literature (CINAHL). A search strategy was developed by expanding upon keywords and combining these with Boolean operators, using the assistance of the Dudley Knowledge Library services. The search strategy used for MEDLINE is presented in Supplementary Figure 1, which was adapted appropriately to search the additional databases.

As few relevant articles relating to SARS-CoV-2 were identified, we deviated from the original protocol search strategy, extending the MEDLINE search to include more terms relating to Long COVID symptoms such as dyspnoea, fever or breathlessness which were noted in our previously published review as the most common symptoms associated with Long COVID^6^ These searches were then supplemented by review of the COVID-NMA database,^18^ the Living Evidence on COVID-19 database^19^ and the first 500 references on both MedRxiv and Google Scholar. However, we did not search for conference or symposia proceedings.

Backward and forward citation searching of the selected studies was conducted to identify further relevant studies. Where possible, authors of protocol studies, which described a suitable trial for inclusion, were contacted to ascertain whether the findings for their trial were completed.

### Study selection

The study selection and quality appraisal stages of the review were facilitated using the online review software Covidence.^20^ From the search results, duplicates were removed, and then titles and abstracts were randomly allocated for screening by at least two independent reviewers. Screening criteria were defined according to whether studies: (i) included patients with a diagnosis of a PVS and (ii) documented information about non-pharmacological treatments. Any disagreements were resolved by a third independent reviewer (JSC and/or OLA).

Following the initial study screening, full-text articles were obtained and assessed according to the full inclusion and exclusion criteria. At this stage, all studies were again reviewed by two independent reviewers. Any disagreements were resolved by two additional independent reviewers (JSC and/or OLA).

### Data extraction and quality appraisal

Relevant data were extracted from the included articles using a predefined extraction criterion. The data extraction was initially piloted by two reviewers (KB and NSW). The data from each study were independently extracted by at least two authors (KB, NSW and JSC), with JSC deciding on the consensus where disparities occurred.

The extracted data included: Title, authors, country where study took place, setting, publication year, study recruitment and follow up dates, study design, participant inclusion and exclusion criteria, sample size, baseline population characteristics for age, sex and ethnicity, virus being studied, PVS definition, reported symptoms, health outcomes reported for those with PVS, intervention description and number of patients allocated to the intervention arm, comparator description and number of patients allocated to the comparator arm, outcome of interest (including method of measurement), and description of main findings.

Included studies were appraised using the Cochrane Risk of Bias tool (RoB-2)^21^, which is also the default appraisal tool in Covidence. The risk of bias was examined by at least two reviewers (KB, NSW, SCR, CM, JSC and NB). Where there were disparities, NB adjudicated between the reviewers.

### Narrative synthesis

We anticipated considerable heterogeneity in the definition of PVS would limit our ability to undertake any pooled analysis or assess for publication bias. We therefore planned a narrative analysis describing the findings, characteristics, and outcomes of the included studies.^22^

### Patient and public involvement

The Patient and Public Involvement and Engagement (PPIE) Group for the Therapies for Long COVID Study^23^ was involved in the co-development of the research question, helped determine the review’s scope and commented on the findings. The patient and public contribution was recorded using the GRIPP-2 short form checklist (supplementary table 2).^24^

## Results

### Description of studies

The study selection process is outlined in Figure 1. Initially, 11,164 results were retrieved, of which 10,631 were identified as non-duplicate. Of these, 10,564 were excluded following title and abstract screening (reviewers at this stage included JSC, OLA, KB, NSK, DH, GT, SCR, RH, SHE, CM and SH), resulting in 67 eligible studies for full-text analysis, of which, 5 were suitable for inclusion. The 67 studies excluded at full-text analysis were due to either incorrect study design, incorrect patient population, or a pharmacological intervention.

**Figure 1.**
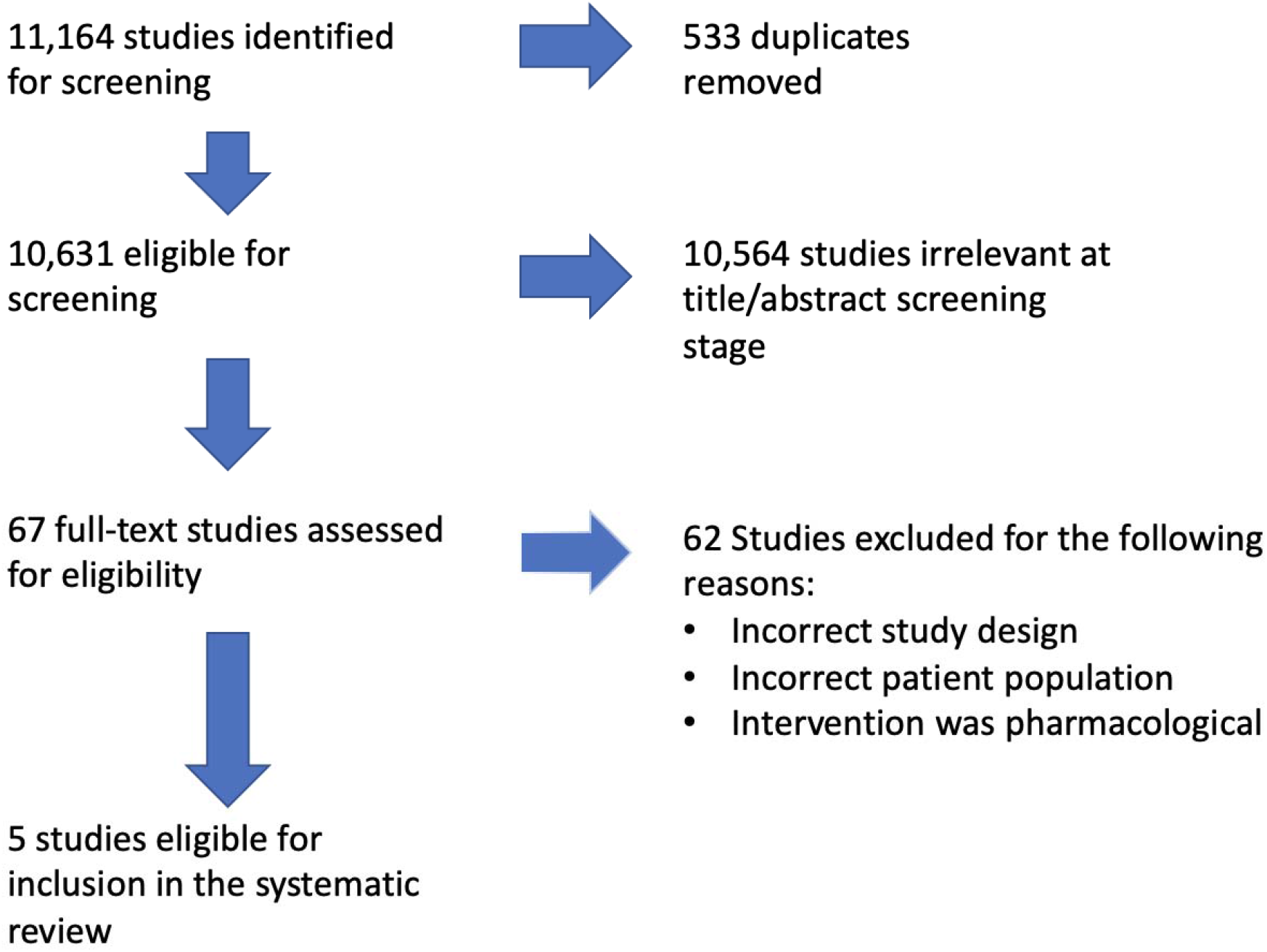
PRISMA Study selection

Our search strategy yielded 11,164 results, which was reduced to 10,631 results after de-duplication. Following, title and abstract screening (reviewers at this stage included JSC, OLA, KB, NSK, DH, GT, SCR, RH, SHE, CM and SH), 67 articles were included for full text review. 62 studies were then excluded (reviewers at this stage included DH, RH, KB, NSW, JSC, SH, OLA) due to either having; 1) an incorrect study design (i.e. not a randomised controlled trial or observational study with a control group), 2) an incorrect patient population (patients did not have symptoms which were either chronic or persistent enough to fulfil our criteria for PVS) and 3) a pharmacological intervention.^25–86^ Following the full text screening, five papers were deemed appropriate to include in the narrative synthesis (further details in figure 1).^13,87–90^

The five studies included were RCTs, further details of their relevant characteristics are described in Table 1.^13,87–90^ The studies were conducted in high income and upper middle-income countries, including China (n=1), Norway (n=1), and Brazil (n=3). Adults (aged 18 years and above) formed the study cohort in four of the RCTs, with children and young people (aged 12-20 years) forming the study group for the RCT conducted in Norway. No studies reported ethnicity, one study reported an intervention relevant to patients with exposure to SARS CoV-2, another relevant to patients experiencing EBV, and three for patients with Chikungunya virus. It is important to note that none of the studies explicitly were designed to capture the full range of symptoms experienced by the participants as part of their PVS. However, the primary symptoms captured were dyspnoea, arthralgia, fatigue (including post-exertional malaise) and general pain which were often used outcome markers. Additionally, as secondary outcomes, many of the studies also captured aggregated data from surveys capturing health related quality of life.

**Table 1.**
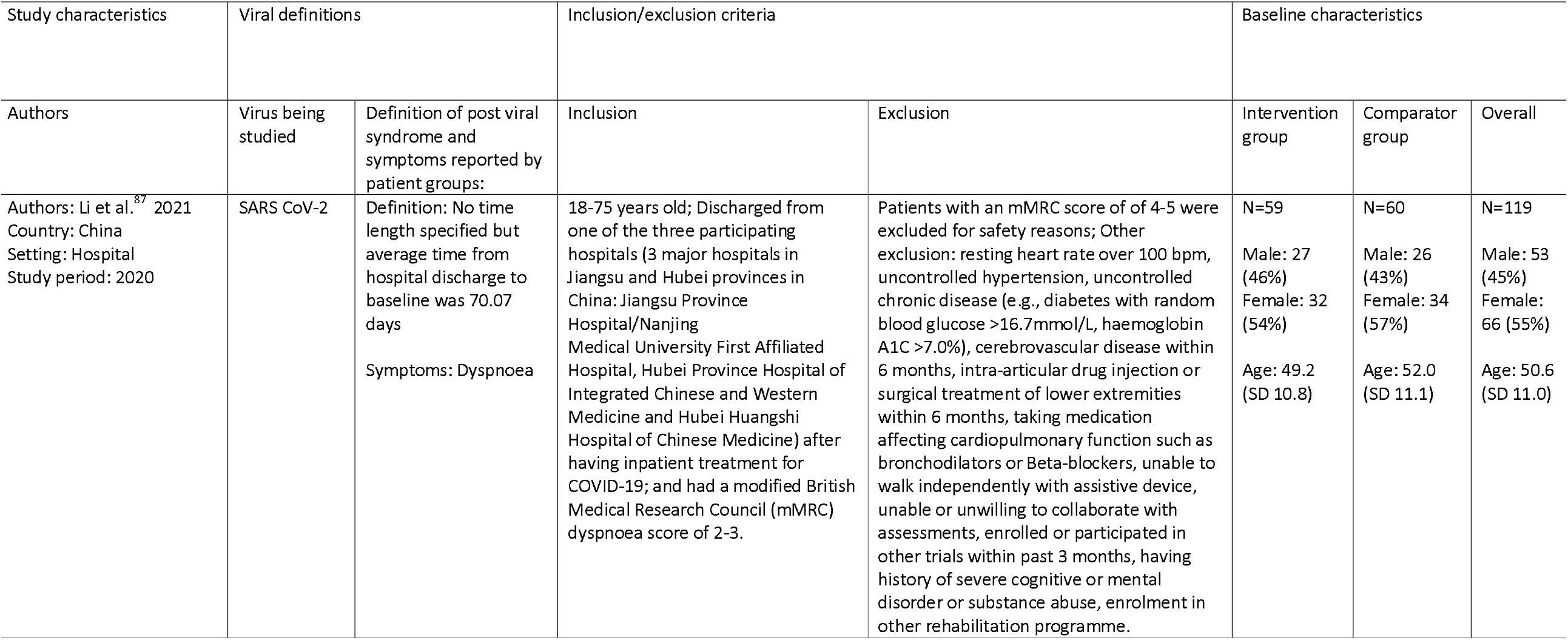
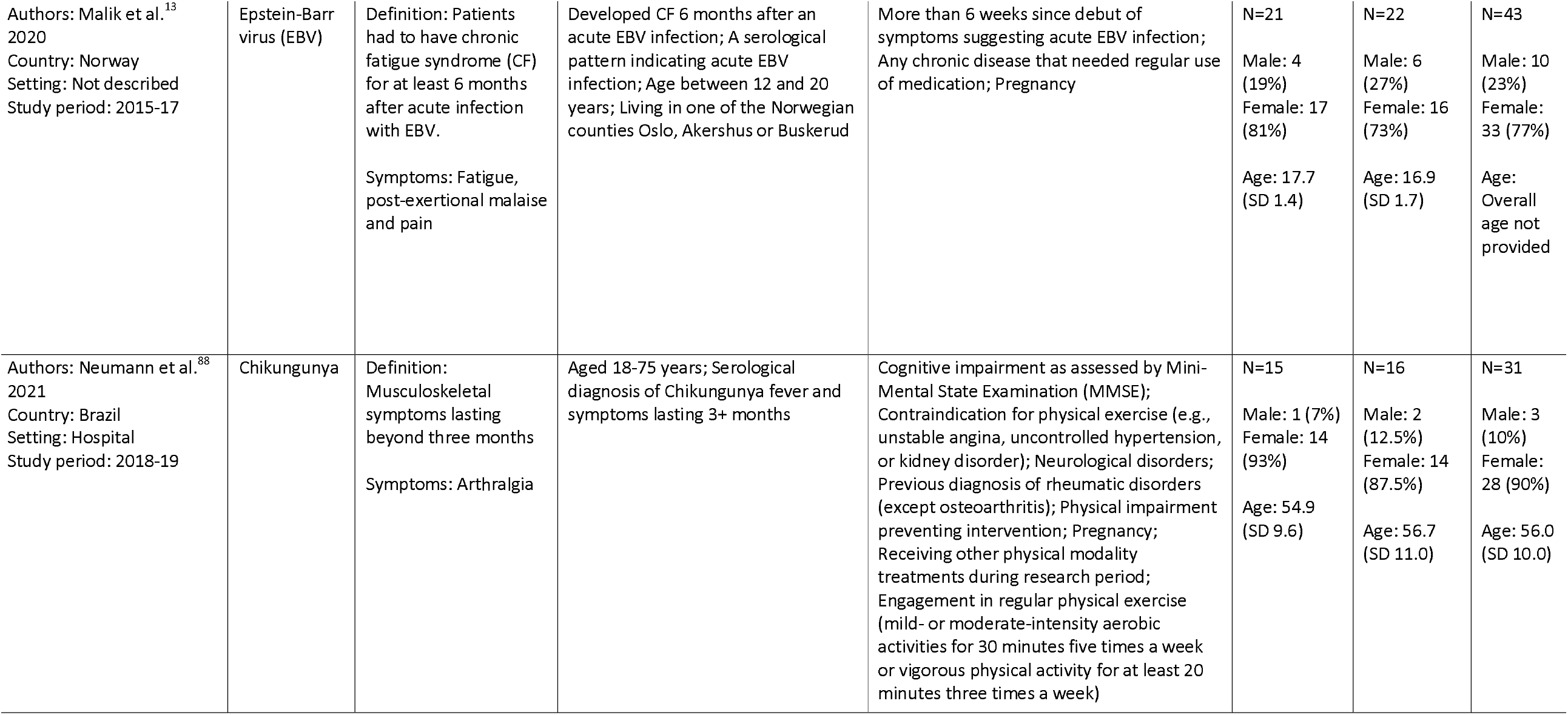
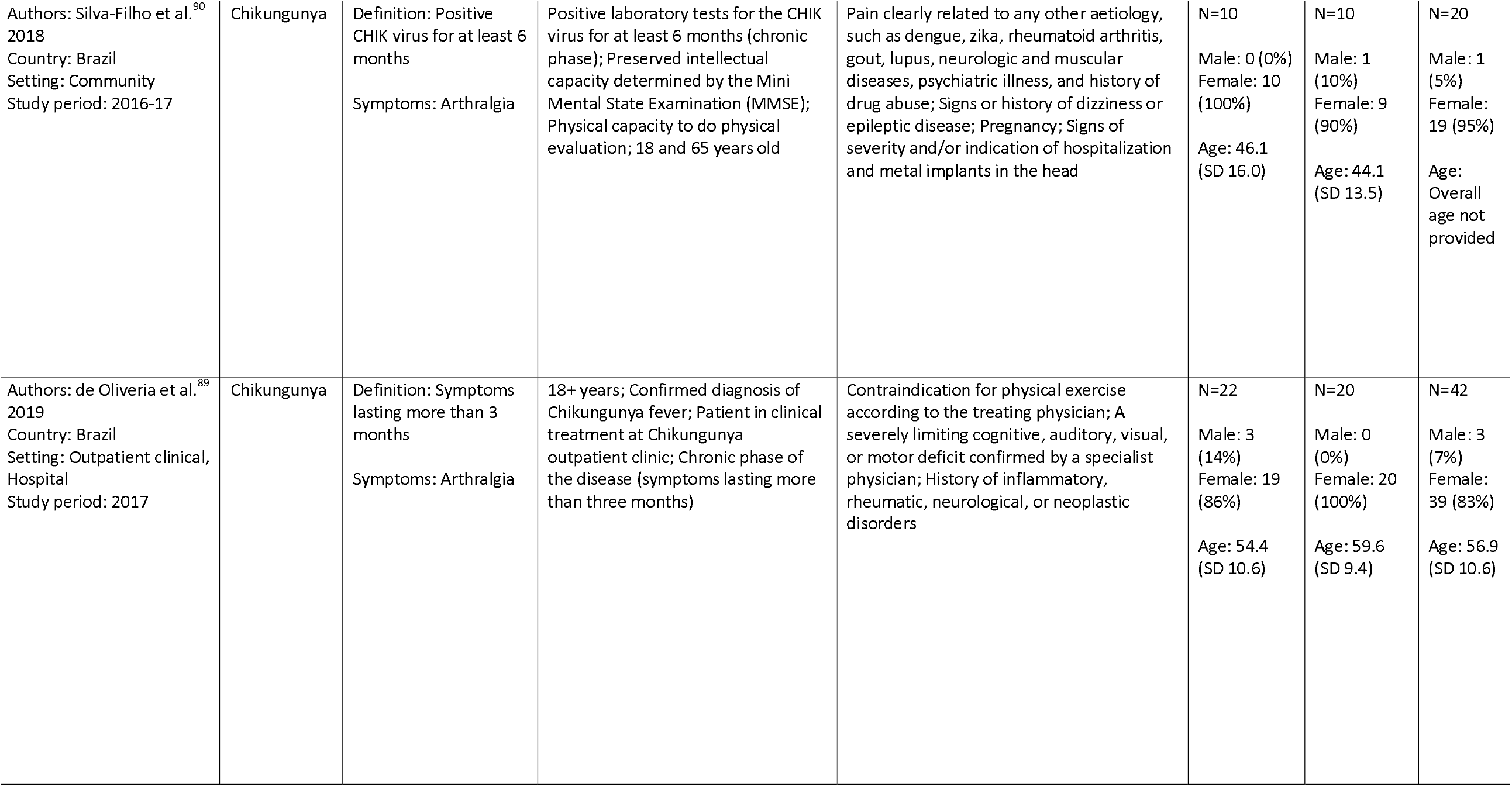
Study Characteristics

**Table 2.** Intervention design, outcome measures and key findings of interventions

| Authors | Description of intervention and comparator | Outcome of interest (including method of measurement) | Description of key findings |
| --- | --- | --- | --- |
| Li et al.<sup>87</sup> | Intervention: Telerehabilitation programme in post-discharge COVID-19 patients (TERECO) is an unsupervised home-based 6-week exercise programme comprising breathing control and thoracic expansion, aerobic exercise, and lower limb muscle strength (LMS) exercise, delivered via smartphone, and remotely monitored with heart rate telemetry.<br>Comparator: One off short educational instruction at baseline | Primary outcome:<br>- Functional exercise capacity - 6 min walking test (6MWT) at 6 weeks post treatment<br>Secondary outcomes:<br>- Functional exercise capacity – 6MWT at 28 weeks follow up<br>- Lower limb muscle strength (LMS) – Static squat test<br>- Pulmonary function – Spirometry<br>- Health-related quality of life (HRQOL) – Short Form Health Survey (SF-12)<br>- Perceived dyspnoea – modified Medical Research Council (mMRC) score | Primary outcome:<br>The mean 6MWD in the control group increased by 17.1 m (SD 63.9) from baseline to post-treatment assessment, whereas 6MWD in the TERECO group improved by 80.2 m (SD 74.7). The adjusted between-group difference in change in 6MWD from baseline (treatment effect) was 65.5 m (95% CI 43.8 to 87.1; $p < 0.001$).<br>Secondary outcomes:<br>- Functional exercise capacity- Estimated 68.6 m (95% CI 46.4 to 90.9; $p < 0.001$) treatment effect difference at follow up<br>- LMS- Improved treatment effect difference 20.1s in squat position (95% CI 12.3 to 27.9; $p < 0.001$) post-treatment, and 22.2s (95% CI 14.2 to 30.2; $p < 0.001$) at follow-up<br>- Pulmonary function- Improvements were seen in both groups. No group differences were found apart from an improvement in maximum voluntary ventilation in the intervention group at post treatment.<br>- HRQOL- Statistically significant improvements in physical component score but not in mental component score.<br>- Perceived dyspnoea- Improvements in mMRC at post-treatment but not at follow up. |
| Malik et al.<sup>13</sup> | Intervention: The intervention consisted of a 10-week mental training programme. The patients had one introductory session followed by nine individual therapy sessions (one per week) for 1.5 hours and related homework, combining elements from CBT and music therapy. Of the nine therapy sessions, four were given by a music therapist and five were given by a cognitive therapist.<br>Comparator: Care as usual- As reported by the authors “neither General Practitioners | Physical activity:<br>- Steps per day – Measured used activPAL accelerometer device<br>Symptoms<br>- Subjective experience of physical & mental fatigue - Chalder Fatigue Questionnaire [CFQ]<br>- Post exertional malaise – Patients were asked ‘how often do you experience more fatigue the day after an exertion?’ on a Likert scale<br>- Pain severity - Brief Pain Inventory<br>- Insomnia and other sleep disturbances - Karolinska Sleep Questionnaire | There were no statistically significant differences between the two groups for any outcomes.<br>Primary outcome:<br>The mean number of steps per day decreased in the treatment group from 3 months post baseline (7217) to 5680 at 15 months post baseline. A decrease was seen in the control group too from 8515 at 3 months post baseline to 7587 at 15 months. The difference between the two groups was not statistically significant: difference (95% CI) = -1298 (-4874 to 2278)). |
|  | or paediatricians in Norway schedule appointment with postinfectious CF patients unless they have strongly reduced physical function. Thus, 'care as usual' implies that the relevant individuals would not receive any healthcare for their CF condition in the follow-up period apart from the follow-up visits in the present study." | - Depression and Anxiety - Hospital Anxiety and Depression Scale<br>- Quality of life - Paediatric Quality of Life Inventory)<br>- Disability related to everyday activities - Functional Disability Inventory<br>- Adverse effects - self-reported |  |
| Neumann et al.<sup>88</sup> | Intervention: The intervention group underwent resistance exercises with elastic bands (24 sessions over 12 weeks) supervised by a physical therapist. The exercise begins with a 5-minute warm up on a stationary bike with no load, followed by resistance exercises for muscle groups that stabilize the shoulders, elbows, wrists, knees and ankles.<br>Comparator: Participants maintained their usual care of treatment and only received phone calls to monitor their symptoms | Primary outcome:<br>- Physical function: Assessed using:<br>- 30-second Chair Stand Test (30-s CST)<br>- 40-meter Fast-paced Walk Test (40m FPWT)<br>- 4-step Stair Climb Power Test (4SCPT)<br>- Disabilities of the arm, shoulder, hand (DASH) questionnaire<br>Secondary outcomes:<br>- Pain intensity- Visual Analogue Scale (VAS) and Disease Activity Score 28 (DAS28)<br>- Quality of life- Medical Outcomes Study 36-Item Short-Form Health Survey (SF36)<sup>91</sup><br>- Patient's global impression – Patient's Global Impression of Change (PGIC) scale (Portuguese version) only asked to intervention group | Primary outcomes:<br>There was a significant improvement between the groups on the 30s CST, with the resisted exercise group improving their performance compared to the control group (p=0.04, d=0.39) at 12 weeks follow up. However, there was no significant improvement in the FPWT, 4SCPT or DASH.<br>Secondary outcomes:<br>- Pain intensity - Reduction in pain intensity (P=0.01; d=-0.83) but no change in painful joints count<br>- SF36- No significant differences<br>- PGIC- 70% of participants in intervention group reported improvement on PGIC scale<br>- No adverse effects were reported in the intervention group |
| Silva-Filho et al.<sup>90</sup> | Intervention: Patients randomised to intervention arm were given transcranial direct current stimulation (tDCS) arm where they experienced a constant current of 2mA for 20 minutes.<br>Comparator: Sham-tDCS was performed on 5 consecutive days with electrodes placed on the same position, and a constant | Primary outcome:<br>- Pain intensity – Assessed using the visual analogue scale (VAS) at baseline, after day 5 of tDCS and at one week follow up.<br>Secondary outcomes:<br>- Pain characteristics – McGill Pain questionnaire and brief pain inventory ((BPI) short form)<br>- Physical function: Assessed using: | Primary outcome:<br>There was a statistically significant improvement in VAS in the tDCS group.<br>Secondary outcomes:<br>- Pain characteristics- There were significant improvements in both the McGill pain questionnaire and BPI in the tDCS group<br>- Physical function- There were no statistically significant differences |
|  | current of 2mA was delivered only for 30s (10-s ramp-up) of the 20min. | - Hand grip test- Hydraulic hand dynamometer<br>- 30-second chair stand test<br>- Upper limb flexion strength- 30-second arm curl test<br>- Physical flexibility: Assessed using:<br>- Chair sit and reach test<br>- Scratch flexibility test<br>- Quality of life-(SF-36) | - Physical flexibility- No improvement in chair sit and reach test but improvement in scratch flexibility test<br>- Quality of life- No difference in SF-36 between the groups |
| de Oliveira et al.<sup>89</sup> | Intervention: The intervention group received 24 sessions of Pilates over a 12-week period. Patients had 2 sessions per week for 50 mins per session, and of light-to-moderate intensity (increasing the number of repetitions, starting with 6 and increasing to 12 repetitions). The exercises involved coordination, strength, flexibility, and balance.<br>Comparator: Usual follow-up at the Chikungunya outpatient clinic, with standard clinical care for the treatment of the disease. | Primary outcome:<br>- Pain intensity- Visual analogue scale (VAS)<br>Secondary outcomes:<br>- Joint range of motion – Joint goniometry<br>- Functioning – Health Assessment Questionnaire<br>- Quality of life – Short-Form Health Survey (SF-12)<sup>92</sup> | Regarding the primary and secondary outcomes, in the intragroup analysis, a significant improvement was observed in all parameters after 24 Pilates sessions (week 12) in relation to the baseline (week 0), but the same was not observed in the control group.<br>The relative risk of an individual having been treated with Pilates and having decreased pain (measured by VAS) was 0.48 (95% CI= 0.28-0.82, P<0.0001) with a number needed to treat two patients (95% CI 7-2, P<0.0001). |

### Risk of bias in included studies

The methodological risk of bias of the included RCTs was generally low Figure 2. However, none of the studies blinded participants and personnel, meaning it was not possible to rule out the risk of a placebo effect or performance bias. Supplementary table 3 contains a narrative description rationalising the risk of bias assessment for each paper from the consensus reviewer (NB).

**Figure 2.**
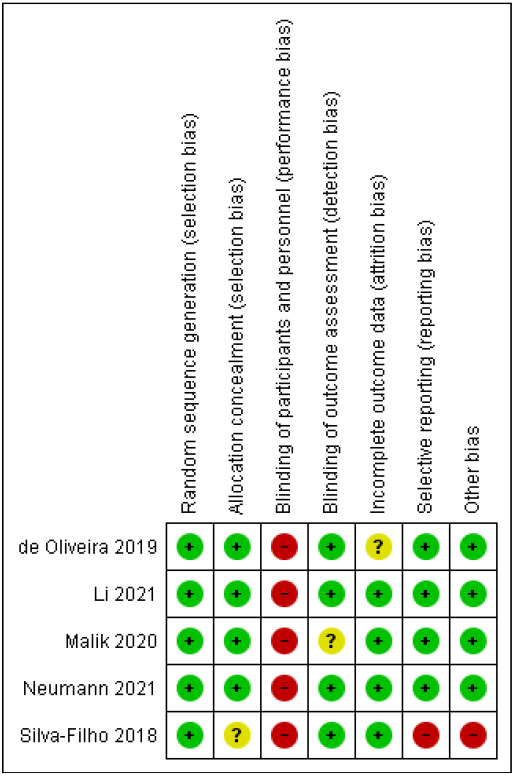
Risk of bias

### Effects of interventions

Due to the heterogeneity in terms of viral exposure (SARS CoV-2, EBV or Chikungunya virus), experienced symptoms (including dyspnoea, fatigue, malaise, pain, insomnia/sleep disturbances, depression/anxiety, and arthralgia) and intervention description, the data could not be combined to perform a meta-analysis. Instead, we have narratively outlined the efficacy of the five included interventions. Table 3 summarises the intervention design, outcome measures and key findings in each of the included trials.

#### Tele-rehabilitation

We identified one published trial describing an intervention designed to support patients with prolonged moderate shortness of breath following exposure to SARS CoV-2.^87^ The telerehabilitation programme in post-discharge COVID-19 patients (TERECO) was an unsupervised home-based 6-week exercise programme comprising breathing control and thoracic expansion, aerobic exercise, and lower limb muscle strength (LMS) exercise, delivered via smartphone, and remotely monitored with heart rate telemetry. Patients were recruited from three major hospitals from Jiangsu and Hubei provinces, China and block randomised to either the intervention group or a control group consisting of those who received a 10-minute standardised instruction from a physiotherapist and a written information sheet containing these instructions. Both groups were advised to maintain normal activities, adhere to a healthy diet, and follow existing public health measures (handwashing, masks, and social distancing).

In this trial, the TERECO programme was superior to the control group in the primary outcome measure at the 6 week follow up point with regards to functional exercise capacity measured using the 6-minute walking distance (6MWD) test. The mean 6MWD in the control group increased by 17.1 m (SD 63.9) from baseline to post-treatment assessment, whereas in the TERECO group, it improved by 80.2 m (SD 74.7). The adjusted between-group difference in change in 6MWD from baseline (treatment effect) was 65.5 m (95% CI 43.8 to 87.1; p<0.001). The TERECO group maintained this benefit at 28 weeks follow up. They also demonstrated improvements in lower limb muscle strength and physical components of the SF-12. However, there were no improvements in spirometry measured pulmonary function or mental health components of the SF-12. There were improvements in perceived dyspnoea measured using the modified medical research council (mMRC) score at 6 weeks post treatment, but this improvement was not statistically significantly different at 28 weeks follow up.

#### Music therapy in combination with cognitive behavioural therapy

Malik et al. explored the effectiveness of a 10-week mental health training programme consisting of a combination of cognitive behavioural therapy and music therapy.^13^ They recruited patients who met their eligibility criteria from the Chronic Fatigue following acute Epstein-Barr Virus infection in Adolescents (CEBA) prospective cohort study based in Norway.^93^ Patients were then randomly allocated to either mental health training programme or ‘care as usual.’ Malik et al. describe that in Norway, neither General Practitioners nor Paediatricians schedule appointments with postinfectious chronic fatigue patients unless they have strongly reduced physical function. Therefore, ‘care as usual’ implies that the relevant individuals would not receive any healthcare for their CF condition in the follow-up period. The outcomes of interest were the subjective experience of physical and mental fatigue, post-exertional malaise, pain severity, insomnia and sleep disturbances, depression and anxiety, quality of life, disability related to everyday activities, steps per day and rates of adverse experiences. The authors found there were no statistically significant differences in any of the outcomes between the two groups. However, they noted that the study was underpowered and recommended the trial should be treated as an exploratory study as sufficient power is needed to assess the effectiveness of this treatment modality.^13^

#### Resistance exercises

Neumann et al. undertook an RCT examining the effectiveness of a resistance exercise programme for patients experiencing prolonged musculoskeletal symptoms who had been exposed to the Chikungunya virus and attended a rheumatology outpatient clinic.^88^ Patients randomly allocated to the intervention arm undertook a resistance exercise programme with elastic bands over a 12-week period with a physical therapist aiming to strengthen muscle groups that stabilise the main functional joint groups affected by Chikungunya disease. Those in the comparator group maintained their usual treatment pathway prescribed by their rheumatologist and received two weekly telephone calls to assess their symptoms. The primary outcome of interest was physical function which was assessed by the 1) 30 second chair stand test (CST), 2) 40-metre fast pace walk test (FPWT), 3) 4-step stair climb power test (4SCPT) and 4) disabilities of arm, shoulder, hand (DASH) questionnaire.

There was a significant improvement in the intervention group on the 30s CST (p=0.04, d=0.39) at 12 weeks follow up compared to the control group. However, there was no significant improvement in the FPWT, 4SCPT or DASH. There was a reduction in pain intensity (secondary outcome) measured using the visual analogue scale, but there was no difference in the disease activity score 28 measuring painful joints or SF-36 measuring quality of life. The patient’s global impression of change (PGIC) score is a validated instrument for measurement of perception of changing health status and treatment satisfaction in patients with chronic musculoskeletal pain and was measured only in the intervention arm, with 70% reporting improvements.

#### Pilates

De Oliveira et al. evaluated the effect of Pilates on pain reduction, improvement of joint function, and quality of life in patients with chronic Chikungunya fever.^89^ Patients were randomly allocated to either a 12 week Pilates exercise programme (two sessions per week for 50 minutes per session at light to moderate intensity) or a control group who did not undergo Pilates. All patients continued to receive usual follow up care at the Chikungunya outpatient clinic. The primary outcome was pain intensity which was assessed using a visual analogue scale on a 0 to 10 scale on a 10cm line. Secondary outcomes were joint range of motion, function, and quality of life. By the 12 week follow up point those who underwent the Pilates intervention compared to the control group experienced a statistically significant improvement in pain intensity with a relative risk of a person being treated with Pilates and having decreased pain being 0.48 (95% CI 0.28-0.82), with a number needed to treat being two patients (95% CI 7-2). All secondary outcome measures showed statistically significant improvements compared to the control group.

#### Neuromodulation

Silva-Filho et al. evaluated the effects of neuromodulation in patients with Chikungunya virus on reduction of joint pain.^90^ Transcranial direct current stimulation (tDCS) is a battery-powered non-invasive neuromodulation technique in which low amplitude direct current is conducted to the cerebral cortex.^90,94^ Patients were either randomised to the tDCS arm where they experienced a constant current of 2mA for 20 minutes or the sham tDCS group where electrodes were placed on the same position but only experienced a constant 2mA current for 30 seconds. The primary outcome, pain intensity, was captured using visual analogue scales by a researcher at baseline, after day 1 and 5 of tDCS and one week follow up.

The VAS scores did not differ substantially between the Active-tDCS and Sham-control group at any of the four time points considered. However, in the Friendman test, a statistically significant decrease in pain over time was found only in the Active-tDCS (p < 0.05, Friedman), and not in the Sham-control group. There were also improvements in secondary outcome measures relating to pain characteristics (measured using the McGill pain questionnaire, brief pain inventory and in the scratch flexibility test). However, there were no improvements in physical function (assessed using the hand group test, 30 second chair stand test, 30 second arm curl test), flexibility assessed using the chair sit and reach test or health-related quality of life measured using the SF-36.

## Discussion

To our knowledge, this is the first systematic review reporting the effectiveness of non-pharmacological treatments evaluated through RCTs for patients experiencing PVS including from viruses other than COVID-19 and observational studies for patients with Long COVID. We sought to synthesise knowledge that can be used to support service planning for the management of patients with Long COVID, in light of the COVID-19 pandemic. We identified five relevant trials that described five distinct types of interventions to support those experiencing chronic PVS-related symptoms. Four of the five (tele-rehabilitation, resistance exercises, Pilates and neuromodulation) interventions reported statistically significant benefits in their primary outcomes, whereas music therapy combined with CBT did not demonstrate significant improvements in any of the measured outcomes. However, it should be noted that the study exploring music therapy with CBT was not adequately powered and further evaluation is needed to assess its efficacy. As seen in this review, the evaluation of non-pharmacological treatments for patients experiencing PVS including Long COVID are limited, and clinical trials are urgently needed to evaluate further therapies and confirm existing findings.

As we identified, there are limited trials of interventions designed to treat these common and persistent symptoms (fatigue being the most commonly reported in patients with Long COVID).^5,6^ Based on our study inclusion criteria, we were unable to find any trials conducted to support patients with ongoing fatigue symptoms following confirmed viral exposure. However, a recent systematic review with broader inclusion criteria, which included both pharmacological and non-pharmacological interventions to support patients with unexplained chronic fatigue syndrome/fibromyalgia, identified forty relevant trials.^14^ Despite the number of these trials, the authors of that review found that relatively few approaches were effective in managing fatigue and of those included, the existing evidence only applied to a narrow range of people with fatigue, a relatively homogeneous group of patients in an age group between 45–55 years, which is not representative of the whole patient cohort thought to experience Long COVID in the UK and elsewhere.^5,14^

Although we did not find any suitable interventions to support patients with virus related fatigue, the TERECO intervention was designed for patients following SARS-CoV-2 exposure and an mMRC score of 2-3 which indicates moderate dyspnoea, the second most commonly reported symptom of Long COVID.^5,6,87^ While this study did not lead to a prolonged improvement in perceived dyspnoea, it showed improvements in the primary outcome which was physical function.^87^ Notably, the TERECO intervention is a multi-component rehabilitation intervention which due to its remote nature can be delivered at scale to support patients who are severely limited by their post viral symptoms and unable to attend a clinic in person as well as during periods of public health restrictions.^87^ A recent review of rehabilitation methods delivered remotely to support physical function has demonstrated that telerehabilitation approaches could be comparable with in-person rehabilitation for a variety of chronic condition management programmes, including cardiac and pulmonary rehabilitation.^95^

Although, the TERECO intervention was the only published trial examining a pulmonary rehabilitation approach to support patients with Long COVID, pulmonary rehabilitation has been thoroughly evaluated in other settings such as in patients with chronic obstructive pulmonary disease, demonstrating consistent positive outcomes in terms of dyspnoea, fatigue and quality of life.^96^ Non-controlled studies and those in populations without continuous symptoms in line with our definition of Long COVID, suggest that pulmonary rehabilitation techniques are beneficial in patients who have experienced COVID-19.^26,27,30,38,43,47,53,54,56,59,60,63,66,70,78,85^ Pulmonary rehabilitation is likely to benefit those experiencing dyspnoea following COVID-19 but further trial evidence is needed to support the appropriateness, effectiveness and cost-effectiveness in those with symptoms lasting at least 12 weeks. Such evidence might be provided by ongoing RCTs designed for this purpose, such as the Rehabilitation Exercise and Psychological Support after COVID-19 Infection (REGAIN)trial which is evaluating the effectiveness of a rehabilitation programme for adults with ongoing COVID-19 sequelae for more than three months after hospital discharge.^97^

The other interventions (Pilates, resistance exercises and neuromodulation), which we identified to be effective for patients experiencing PVS were all examined in patient populations with Chikungunya virus exposure.^88–90^ All of these interventions identified benefits in patients’ experiences and perceptions of pain intensity, in particular arthralgia, which is another common symptom experienced by patients with Long COVID.^6,88–90^ Although, these have not yet been formally evaluated in patients with Long COVID, they may improve the symptom burden in this patient population. For example, case reports have demonstrated benefits of neuromodulation particularly in the management of the mental health effects of COVID-19 and consequently, there are RCTs underway such as the Symptoms, Trajectory, Inequalities and Management: Understanding Long-COVID to Address and Transform Existing Integrated Care Pathways (STIMULATE-ICP) and Home-based Brain Stimulation Treatment for Post-acute Sequelae of COVID-19 (PASC) trials to examine the efficacy of this therapy modality in patients with Long COVID.^98–102^

A key challenge with these interventions (Pilates, resistance exercises and neuromodulation) relates to their scalability, which is particularly important when considering the scale of the public health burden in a context of limited health service capacity. However, these therapies were not assessed in trials for home use. Additionally, relating to Pilates and resistance exercises, NICE recently recommended against the use of graded exercise therapies in supporting patients with Myalgic Encephalitis (ME) and Chronic Fatigue Syndrome (CFS).^103^ Although, the aetiology of ME/CFS remains under investigation, patients with these conditions experience ongoing fatigue similar to Long COVID. Through expert consultation, it was deemed that people experiencing such symptoms should undertake therapy options where they remain within their energy limits and care should be given to undertake activities that do not worsen symptoms.^103^ Therefore, before widespread adoption of exercise-based therapies such as Pilates or resistance exercises is considered for patients with Long COVID, further research is needed to identify which patients are most likely to benefit from these therapies.

Being reported in line with the PRISMA statement, our systematic review has several strengths such as the inclusion of a comprehensive search strategy encompassing pre-print databases in addition to peer reviewed articles. The continued PPI support for this paper (as highlighted in Supplementary Table 2) was a particular strength of the paper as we were able to formulate the review design with input from people with lived experience of Long COVID.

However, there were some limitations to the study design and some minor deviations from the published protocol. For example, the scope of the review had increased after identifying few relevant studies in the initial search which only included patients with PVS in RCT settings. Due to the nature of our search strategy and study aim, we did not capture evidence on interventions which were examined using non-trialled and potentially less robust methods. However, we acknowledge that some such studies may indeed provide information relevant to the affected patient population and clinicians involved in the management of Long COVID. An additional limitation we noted during our review was the limited nature of symptom burden reporting in the studies included. There were two challenges posed in respect to this 1) the studies did not often describe the breadth of symptoms experienced by participants. The variety and combination of symptoms experienced by participants may impact on the effectiveness of the interventions and 2) the studies included did not have standardised methods of reporting symptom related outcome measures which also limited comparability when assessing effectiveness. As such, we recommend that future research in this area is undertaken to develop standardised outcome measurement tools to facilitate evaluation of relevant interventions pertaining to supporting patients with PVS. With the support of patients, public and clinicians, we have developed a symptom burden questionnaire for Long COVID (SBQ-LC) which could fulfil this need in contemporary studies.^104^

## Conclusions

The aim of this systematic review was to identify the existing evidence base for non-pharmacological treatments which can be delivered to support patients with PVS, including Long COVID. The findings of this review identified few treatment modalities which have been evaluated to determine their application to patients with Long COVID. Considering the extensive public health burden of Long COVID, there is an urgent need for further trials to evaluate supportive interventions for chronic symptoms following SARS-CoV-2 exposure, as well as other viral pathogens, and to build upon the knowledge base across overlapping symptoms.

## Data Availability

All data produced in the present work are contained in the manuscript

## Funding and acknowledgments

This work is independent research jointly funded by the National Institute for Health and Care Research (NIHR) and UK Research and Innovation (UKRI) (Therapies for Long COVID in non-hospitalised individuals: From symptoms, patient reported outcomes and immunology to targeted therapies (The TLC Study), COV-LT-0013). The views expressed in this publication are those of the author(s) and not necessarily those of the NIHR, the Department of Health and Social Care or UKRI. The authors are grateful to Dudley Knowledge Library services (Carl Cross and Abimola Alayo) who supported the development and execution of the search strategy.

## Notes

**Conflict of interest for this study:** There are no conflicts of interest among the authors relevant to this study.

### Competing Interest Statement

The authors have declared no competing interest.

### Clinical Protocols

https://www.crd.york.ac.uk/prospero/display_record.php?ID=CRD42021282074

## References

1 Johns Hopkins University. COVID-19 Map - Johns Hopkins Coronavirus Resource Center. https://coronavirus.jhu.edu/map.html (accessed 7 Jun2022).

2 Haas EJ, Angulo FJ, McLaughlin JM, Anis E, Singer SR, Khan F et al. Impact and effectiveness of mRNA BNT162b2 vaccine against SARS-CoV-2 infections and COVID-19 cases, hospitalisations, and deaths following a nationwide vaccination campaign in Israel: an observational study using national surveillance data. Lancet 2021; 397: 1819–1829.

3 Public Health England. Direct and indirect impact of the vaccination programme on COVID-19 infections and mortality. 2021.

4 Gupta S, Cantor J, Simon KI, Bento AI, Wing C, Whaley CM. Vaccinations Against COVID-19 May Have Averted Up To 140,000 Deaths In The United States. Health Aff 2021; 40: 1465–1472.

5 Office for National Statistics. Prevalence of ongoing symptoms following coronavirus (COVID-19) infection in the UK. 2022.https://www.ons.gov.uk/peoplepopulationandcommunity/healthandsocialcare/conditionsanddiseases/bulletins/prevalenceofongoingsymptomsfollowingcoronaviruscovid19infectionintheuk/7april2022 (accessed 11 May2022).

6 Aiyegbusi OL, Hughes SE, Turner G, Rivera SC, McMullan C, Chandan JS et al. Symptoms, complications and management of long COVID: a review. J R Soc Med 2021; 114: 428–442.

7 Perego E, Callard F, Stras L, Melville-Jóhannesson B, Pope R, Alwan NA. Why we need to keep using the patient made term “Long Covid” - The BMJ. 2020.https://blogs.bmj.com/bmj/2020/10/01/why-we-need-to-keep-using-the-patient-made-term-long-covid/ (accessed 7 Jun2022).

8 Long COVID: let patients help define long-lasting COVID symptoms. Nature 2020; 586: 170.

9 National Institute of Health and Care Excellence. COVID-19 rapid guideline: managing the long-term effects of COVID-19 | Guidance | NICE. 2020.https://www.nice.org.uk/guidance/ng188 (accessed 7 Jun2022).

10 Aucott JN, Rebman AW. Long-haul COVID: heed the lessons from other infection-triggered illnesses. Lancet 2021; 397: 967–968.

11 Salamanna F, Veronesi F, Martini L, Landini MP, Fini M. Post-COVID-19 Syndrome: The Persistent Symptoms at the Post-viral Stage of the Disease. A Systematic Review of the Current Data. Front Med 2021; 8: 392.

12 White PD. What Causes Prolonged Fatigue after Infectious Mononucleosis—and Does It Tell Us Anything about Chronic Fatigue Syndrome? J Infect Dis 2007; 196: 4–5.

13 Malik S, Asprusten TT, Pedersen M, Mangersnes J, Trondalen G, Van Roy B et al. Cognitive–behavioural therapy combined with music therapy for chronic fatigue following Epstein-Barr virus infection in adolescents: a feasibility study. BMJ Paediatr Open 2020; 4. doi:10.1136/BMJPO-2019-000620.

14 Fowler-Davis S, Platts K, Thelwell M, Woodward A, Harrop D. A mixed-methods systematic review of post-viral fatigue interventions: Are there lessons for long Covid? PLoS One 2021; 16: e0259533.

15 Page MJ, McKenzie JE, Bossuyt PM, Boutron I, Hoffmann TC, Mulrow CD et al. The PRISMA 2020 statement: an updated guideline for reporting systematic reviews. BMJ 2021; 372. doi:10.1136/BMJ.N71.

16 PROSPERO. Non-pharmacological therapies for post-viral syndromes, including Long COVID: a systematic review and meta-analysis protocol. 2021.https://www.crd.york.ac.uk/prospero/display_record.php?ID=CRD42021282074 (accessed 7 Jun2022).

17 World Health Organization. A clinical case definition of post COVID-19 condition by a Delphi consensus. 2021.https://www.who.int/publications/i/item/WHO-2019-nCoV-Post_COVID-19_condition-Clinical_case_definition-2021.1 (accessed 7 Jun2022).

18 Boutron I, Chaimani A, Meerpohl JJ, Hróbjartsson A, Devane D, Rada G et al. The COVID-NMA Project: Building an Evidence Ecosystem for the COVID-19 Pandemic. Ann Intern Med 2020; 173: 1015–1017.

19 Counotte M, Imeri H, Heron L, Ipekci AM, Low N. Living Evidence on COVID-19. 2020.https://ispmbern.github.io/covid-19/living-review/ (accessed 7 Jun2022).

20 Veritas Health Innovation. Covidence systematic review software..

21 Sterne JAC, Savović J, Page MJ, Elbers RG, Blencowe NS, Boutron I et al. RoB 2: a revised tool for assessing risk of bias in randomised trials. BMJ 2019; 366. doi:10.1136/BMJ.L4898.

22 Guyatt GH, Oxman AD, Vist GE, Kunz R, Falck-Ytter Y, Alonso-Coello P et al. GRADE: an emerging consensus on rating quality of evidence and strength of recommendations. BMJ 2008; 336: 924–926.

23 Haroon S, Nirantharakumar K, Hughes SE, Subramanian A, Aiyegbusi OL, Davies EH et al. Therapies for Long COVID in non-hospitalised individuals: from symptoms, patient-reported outcomes and immunology to targeted therapies (The TLC Study). BMJ Open 2022; 12: e060413.

24 Staniszewska S, Brett J, Simera I, Seers K, Mockford C, Goodlad S et al. GRIPP2 reporting checklists: Tools to improve reporting of patient and public involvement in research. Res Involv Engagem 2017; 3: 1–11.

25 Nguyen HC, Nguyen MH, Do BN, Tran CQ, Nguyen TTP, Pham KM et al. People with Suspected COVID-19 Symptoms Were More Likely Depressed and Had Lower Health-Related Quality of Life: The Potential Benefit of Health Literacy. J Clin Med 2020; 9: 965.

26 Joshi R, Rathi M, Thakur J. Physiotherapy rehabilitation in post-COVID-19 phase: A case report. J Clin Diagnostic Res 2021; 15: YD01–YD02.

27 Daynes E, Gerlis C, Chaplin E, Gardiner N, Singh SJ. Early experiences of rehabilitation for individuals post-COVID to improve fatigue, breathlessness exercise capacity and cognition – A cohort study. Chron Respir Dis 2021; 18: 147997312110156.

28 Vlake JH, Van Bommel J, Wils E-J, Korevaar TIM, Hellemons ME, Schut AFC et al. Effect of intensive care unit-specific virtual reality (ICU-VR) to improve psychological well-being and quality of life in COVID-19 ICU survivors: a study protocol for a multicentre, randomized controlled trial. Trials 2021; 22: 328.

29 Zha L, Xu X, Wang D, Qiao G, Zhuang W, Huang S. Modified rehabilitation exercises for mild cases of COVID-19. Ann Palliat Med 2020; 9: 3100–3106.

30 Zampogna E, Paneroni M, Belli S, Aliani M, Gandolfo A, Visca D et al. Pulmonary Rehabilitation in Patients Recovering from COVID-19. Respiration 2021; 100: 416–422.

31 Reid MR, Mackinnon LT, Drummond PD. The effects of stress management on symptoms of upper respiratory tract infection, secretory immunoglobulin A, and mood in young adults. J Psychosom Res 2001; 51: 721–728.

32 deLima Thomas J, Leiter RE, Abrahm JL, Shameklis JC, Kiser SB, Gelfand SL et al. Development of a Palliative Care Toolkit for the COVID-19 Pandemic. J Pain Symptom Manage 2020; 60: e22–e25.

33 Meeus M, Nijs J, Van Oosterwijck J, Van Alsenoy V, Truijen S. Pain Physiology Education Improves Pain Beliefs in Patients With Chronic Fatigue Syndrome Compared With Pacing and Self-Management Education: A Double-Blind Randomized Controlled Trial. Arch Phys Med Rehabil 2010; 91: 1153–1159.

34 Curci C, Pisano F, Bonacci E, Camozzi DM, Ceravolo C, Bergonzi R et al. Early rehabilitation in post-acute COVID-19 patients: data from an Italian COVID-19 Rehabilitation Unit and proposal of a treatment protocol. Eur J Phys Rehabil Med 2020; 56: 633–641.

35 Vlake JH, van Bommel J, Wils E-J, Bienvenu J, Hellemons ME, Korevaar TIM et al. Intensive Care Unit–Specific Virtual Reality for Critically Ill Patients With COVID-19: Multicenter Randomized Controlled Trial. J Med Internet Res 2022; 24: e32368.

36 Lee K-M, Ko H-J, Lee GH, Kim A-S, Lee D-W. A Well-Structured Follow-Up Program is Required after Recovery from Coronavirus Disease 2019 (COVID-19); Release from Quarantine is Not the End of Treatment. J Clin Med 2021; 10: 2329.

37 Gruzelier J, Champion A, Fox P, Rollin M, McCormack S, Catalan P et al. Individual differences in personality, immunology and mood in patients undergoing self-hypnosis training for the successful treatment of a chronic viral illness, HSV-2. Contemp Hypn 2002; 19: 149–166.

38 Hayden MC, Limbach M, Schuler M, Merkl S, Schwarzl G, Jakab K et al. Effectiveness of a Three-Week Inpatient Pulmonary Rehabilitation Program for Patients after COVID-19: A Prospective Observational Study. Int J Environ Res Public Health 2021; 18: 9001.

39 Khah MD AS, Mortazavian Msc S, Kateb MY, Najafabadi PhD MG, Niyazi MD S. The effect of online mindfulness program on physical pain, stress and depression in the COVID-19 patients: A randomized control trail. J Pain Manag 2021; 14: 57–63.

40 Parizad N, Goli R, Faraji N, Mam-Qaderi M, Mirzaee R, Gharebaghi N et al. Effect of guided imagery on anxiety, muscle pain, and vital signs in patients with COVID-19: A randomized controlled trial. Complement Ther Clin Pract 2021; 43: 101335.

41 Li J, Li X, Jiang J, Xu X, Wu J, Xu Y et al. The Effect of Cognitive Behavioral Therapy on Depression, Anxiety, and Stress in Patients With COVID-19: A Randomized Controlled Trial. Front Psychiatry 2020; 11: 1096.

42 Liu Z, Qiao D, Xu Y, Zhao W, Yang Y, Wen D et al. The Efficacy of Computerized Cognitive Behavioral Therapy for Depressive and Anxiety Symptoms in Patients With COVID-19: Randomized Controlled Trial. J Med Internet Res 2021; 23: e26883.

43 Sun J, Liu J, Li H, Shang C, Li T, Ji W et al. Pulmonary rehabilitation focusing on the regulation of respiratory movement can improve prognosis of severe patients with COVID-19. Ann Palliat Med 2021; 10: 4262272–4264272.

44 Gonzalez-Gerez JJ, Saavedra-Hernandez M, Anarte-Lazo E, Bernal-Utrera C, Perez-Ale M, Rodriguez-Blanco C. Short-Term Effects of a Respiratory Telerehabilitation Program in Confined COVID-19 Patients in the Acute Phase: A Pilot Study. Int J Environ Res Public Health 2021; 18: 7511.

45 Kong X, Kong F, Zheng K, Tang M, Chen Y, Zhou J et al. Effect of Psychological– Behavioral Intervention on the Depression and Anxiety of COVID-19 Patients. Front Psychiatry 2020; 11: 1241.

46 Sotoudeh HG, Alavi SS, Akbari Z, Jannatifard F, Artounian V. The Effect of Brief Crisis Intervention Package on Improving Quality of Life and Mental Health in Patients with COVID-19. Iran J Psychiatry 2020; 15: 205.

47 Champion T, Bauer N. Effects of a structured cardiopulmonary rehabilitation program for individuals recovering from covid-19. Cardiopulm Phys Ther J 2021; 32: e5–e6.

48 Polsky MB, Moraveji N. Post-acute care management of a patient with COVID-19 using remote cardiorespiratory monitoring. Respir Med Case Reports 2021; 33: 101436.

49 Chen JM, Wang ZY, Chen YJ, Ni J. The Application of Eight-Segment Pulmonary Rehabilitation Exercise in People With Coronavirus Disease 2019. Front Physiol 2020; 11: 646.

50 Hui F, Boyle E, Vayda E, Glazier RH. A randomized controlled trial of a multifaceted integrated complementary-alternative therapy for chronic herpes zoster-related pain. Altern Med Rev 2012; 17: 57–68.

51 Ye L, Sun P, Wang T. Acupuncture strategies to tackle post covid-19 psychological and neuropsychiatric disorders. J Chinese Med 2020; : 11–21.

52 Keijmel SP, Delsing CE, Bleijenberg G, van der Meer JWM, Donders RT, Leclercq M et al. Effectiveness of Long-term Doxycycline Treatment and Cognitive-Behavioral Therapy on Fatigue Severity in Patients with Q Fever Fatigue Syndrome (Qure Study): A Randomized Controlled Trial. Clin Infect Dis 2017; 64: 998–1005.

53 Mayer KP, Steele AK, Soper MK, Branton JD, Lusby ML, Kalema AG et al. Physical Therapy Management of an Individual With Post-COVID Syndrome: A Case Report. Phys Ther 2021; 101. doi:10.1093/ptj/pzab098.

54 Dalbosco-Salas M, Torres-Castro R, Rojas Leyton A, Morales Zapata F, Henríquez Salazar E, Espinoza Bastías G et al. Effectiveness of a Primary Care Telerehabilitation Program for Post-COVID-19 Patients: A Feasibility Study. J Clin Med 2021; 10: 4428.

55 Tsiskarishvili N, Tsiskarishvili N, Tsiskarishvili T. [Local phototherapy and pulse current in complex treatment of relapsing herpes simplex]. Georg Med News 2007; : 31–4.

56 Liu K, Zhang W, Yang Y, Zhang J, Li Y, Chen Y. Respiratory rehabilitation in elderly patients with COVID-19: A randomized controlled study. Complement Ther Clin Pract 2020; 39: 101166.

57 Szlamka Z, Kiss M, Bernáth S, Kámán P, Lubani A, Karner O et al. Mental Health Support in the Time of Crisis: Are We Prepared? Experiences With the COVID-19 Counselling Programme in Hungary. Front Psychiatry 2021; 12. doi:10.3389/fpsyt.2021.655211.

58 Teixeira IS, Leal FS, Tateno RY, Palma LF, Campos L. Photobiomodulation therapy and antimicrobial photodynamic therapy for orofacial lesions in patients with COVID-19: A case series. Photodiagnosis Photodyn Ther 2021; 34: 102281.

59 Gloeckl R, Leitl D, Jarosch I, Schneeberger T, Nell C, Stenzel N et al. Benefits of pulmonary rehabilitation in COVID-19: a prospective observational cohort study. ERJ Open Res 2021; 7: 00108–02021.

60 Tang Y, Jiang J, Shen P, Li M, You H, Liu C et al. Liuzijue is a promising exercise option for rehabilitating discharged COVID-19 patients. Medicine (Baltimore) 2021; 100: e24564.

61 Cineka A, Raj J. Dance and Music as a Therapy to Heal Physical and Psychological Pain: An Analytical Study of COVID-19 Patients during Quarantine. Eur J Mol Clin Med 2020; 7: 99–109.

62 Imamura M, Mirisola AR, Ribeiro F de Q, de Pretto LR, Alfieri FM, Delgado VR et al. Rehabilitation of patients after COVID-19 recovery: An experience at the Physical and Rehabilitation Medicine Institute and Lucy Montoro Rehabilitation Institute. Clinics 2021; 76. doi:10.6061/CLINICS/2021/E2804.

63 Pan Hj, Bao Xh, Chen Jy, Feng Y, Kang By, Wang Jx et al. Respiratory rehabilitation assisted by respiratory trainers in patients with coronavirus disease 2019: An analysis of efficacy. Acad J Second Mil Med Univ 2021; 42: 255–260.

64 Wang C-Y, Fang J-Q. [Analysis on therapeutic effect of variable-frequency electroacupuncture combined with herbal-moxa moxibustion for post-zoster neuralgia]. Zhen ci yan jiu = Acupunct Res 2012; 37: 64–6.

65 Zhang XB, Zhang JL, Li MX, Yuan YP, Sun J. Baduanjin exercise can alleviate anxiety and depression of patients with COVID-19 in Square cabin hospital: A cross-sectional survey. Medicine (Baltimore) 2021; 100: e26898.

66 Wootton SL, King M, Alison JA, Mahadev S, Chan ASL. COVID=19 rehabilitation delivered via a telehealth pulmonary rehabilitation model: a case series. Respirol Case Reports 2020; 8: 669.

67 Biondi Situmorang DD. Music Therapy for the Treatment of Patients with COVID-19: Psychopathological Problems Intervention and Well-Being Improvement. Infect Dis Clin Pract 2021; 29: E198.

68 Pal GK, Nanda N, Pal P, Renugasundari M, Hariprasad R, Rajesh DR et al. Asanas in prone posture and slow pranayamic breathing promote early recoveryand prevent post-recovery complications in COVID-19 patients: a preliminary study. Biomedicine 2021; 41: 390–396.

69 Voshaar T, Stais P, Köhler D, Dellweg D. Conservative management of COVID-19 associated hypoxaemia. ERJ Open Res 2021; 7: 00292–02021.

70 Ahmed I, Inam A Bin, Belli S, Ahmad J, Khalil W, Jafar MM. Effectiveness of aerobic exercise training program on cardio-respiratory fitness and quality of life in patients recovered from COVID-19. Eur J Physiother 2021; 33: 1–6.

71 Sanger NS, Kankakee A, Singh R, Gupta A, Shrikhande B, Ganu G. A randomized controlled clinical trial to evaluate the potential of ayurvedic medicines in patients with mild symptoms of COVID-19. Eur J Mol Clin Med 2021; 8: 3017–3027.

72 Olenskaya TL, Nikolayeva AH, Piatsko VV, Azaronak MK, Yukhno YS. Application of altitude chamber adaptation as a component of the medical rehabilitation patients after COVID-19. Profil meditsina 2021; 24: 76.

73 Zhou L, Xie R, Yang X, Zhang S, Li D, Zhang Y et al. Feasibility and Preliminary Results of Effectiveness of Social Media-based Intervention on the Psychological Well-being of Suspected COVID-19 Cases during Quarantine. Can J Psychiatry 2020; 65: 736–738.

74 Schindl A, Neumann R. Low-Intensity Laser Therapy is an Effective Treatment for Recurrent Herpes Simplex Infection. Results from a Randomized Double-Blind Placebo-Controlled Study. J Invest Dermatol 1999; 113: 221–223.

75 Shakerian N, Mofateh R, Saghazadeh A, Rezaei N, Rezaei N. Potential Prophylactic and Therapeutic Effects of Respiratory Physiotherapy for COVID-19. Acta Biomed 2020; 92: e2021020.

76 Kurtaiş Aytür Y. Pulmonary rehabilitation principles in SARS-COV-2 infection (COVID-19): A guideline for the acute and subacute rehabilitation. Turkish J Phys Med Rehabil 2020; 66: 104–120.

77 Hewson-Bower B, Drummond PD. Psychological treatment for recurrent symptoms of colds and flu in children. J Psychosom Res 2001; 51: 369–377.

78 Rodriguez-Blanco C, Gonzalez-Gerez JJ, Bernal-Utrera C, Anarte-Lazo E, Perez-Ale M, Saavedra-Hernandez M. Short-Term Effects of a Conditioning Telerehabilitation Program in Confined Patients Affected by COVID-19 in the Acute Phase. A Pilot Randomized Controlled Trial. Medicina (B Aires) 2021; 57: 684.

79 Jenefer Jerrin R, Theebika S, Panneerselvam P, Venkateswaran S, Manavalan N, Maheshkumar K. Yoga and Naturopathy intervention for reducing anxiety and depression of Covid-19 patients – A pilot study. Clin Epidemiol Glob Heal 2021; 11: 100800.

80 Bickton FM, Chisati E, Rylance J, Morton B. An Improvised Pulmonary Telerehabilitation Program for Postacute COVID-19 Patients Would Be Feasible and Acceptable in a Low-Resource Setting. Am J Phys Med Rehabil 2021; 100: 209–212.

81 Cheng SI. Medical Acupuncture as a Treatment for Novel COVID-19-Related Respiratory Distress: Personal Experience from a Frontline Anesthesiologist. Med Acupunct 2021; 33: 83–85.

82 Pal GK, Nanda N, Renugasundari M, Pal P, Pachegaonkar U. Acute effects of prone asanas and Pal’s pranayama on myalgia, headache, psychological stress and respiratory problems in the COVID-19 patients in the recovery phase. Biomedicine 2020; 40: 526–530.

83 Karthikeyan T. Therapeutic effectiveness of diaphragmatic with costal breathing exercises on C-19 PEFR patients. Intensive Care Med Exp 2021; 9.

84 Song J, Jiang R, Chen N, Qu W, Liu D, Zhang M et al. Self-help cognitive behavioral therapy application for COVID-19-related mental health problems: A longitudinal trial. Asian J Psychiatr 2021; 60: 102656.

85 Gilmutdinova IR, Kolyshenkov VA, Lapickaya KA, Trepova AS, Vasileva VA, Prosvirnin AN et al. Telemedicine platform COVIDREHAB for remote rehabilitation of patients after COVID-19. Eur J Transl Myol 2021; 31. doi:10.4081/ejtm.2021.9783.

86 Lerner D, Garvey K, Arrighi-Allisan A, Filimonov A, Filip P, Liu K et al. Letter to the editor: Study Summary -Randomized Control Trial of Omega-3 Fatty Acid Supplementation for the Treatment of COVID-19 Related Olfactory Dysfunction. Trials 2020; 21: 942.

87 Li J, Xia W, Zhan C, Liu S, Yin Z, Wang J et al. A telerehabilitation programme in post-discharge COVID-19 patients (TERECO): A randomised controlled trial. Thorax 2021. doi:10.1136/THORAXJNL-2021-217382.

88 Neumann IL, de Oliveira DA, de Barros EL, Santos GDS, de Oliveira LS, Duarte AL et al. Resistance exercises improve physical function in chronic Chikungunya fever patients: a randomized controlled trial. Eur J Phys Rehabil Med 2021; 57: 620–629.

89 de Oliveira Bfa, Carvalho Prc, de Souza Holanda As, dos Santos Risb, da Silva Fax, Barros Gwp et al. Pilates method in the treatment of patients with Chikungunya fever: a randomized controlled trial. Clin Rehabil 2019; 33: 1614–1624.

90 Silva-Filho E, Okano AH, Morya E, Albuquerque J, Cacho E, Unal G et al. Neuromodulation treats Chikungunya arthralgia: a randomized controlled trial. Sci Rep 2018; 8. doi:10.1038/S41598-018-34514-4.

91 RAND Corporation. 36-Item Short Form Survey (SF-36). https://www.rand.org/health-care/surveys_tools/mos/36-item-short-form.html (accessed 11 May2022).

92 RAND Corporation. 12-Item Short Form Survey (SF-12). https://www.rand.org/health-care/surveys_tools/mos/12-item-short-form.html (accessed 11 May2022).

93 Pedersen M, Asprusten TT, Godang K, Leegaard TM, Osnes LT, Skovlund E et al. Predictors of chronic fatigue in adolescents six months after acute Epstein-Barr virus infection: A prospective cohort study. Brain Behav Immun 2019; 75: 94–100.

94 Boggio PS, Zaghi S, Fregni F. Modulation of emotions associated with images of human pain using anodal transcranial direct current stimulation (tDCS). Neuropsychologia 2009; 47: 212–217.

95 Seron P, Oliveros MJ, Gutierrez-Arias R, Fuentes-Aspe R, Torres-Castro RC, Merino-Osorio C et al. Effectiveness of Telerehabilitation in Physical Therapy: A Rapid Overview. Phys Ther 2021; 101. doi:10.1093/PTJ/PZAB053.

96 Mccarthy B, Casey D, Devane D, Murphy K, Murphy E, Lacasse Y. Pulmonary rehabilitation for chronic obstructive pulmonary disease. Cochrane Database Syst Rev 2015; 2015. doi:10.1002/14651858.CD003793.PUB3/MEDIA/CDSR/CD003793/IMAGE_N/NCD003793-CMP-004-08.PNG.

97 McGregor G, Sandhu H, Bruce J, Sheehan B, McWilliams D, Yeung J et al. Rehabilitation Exercise and psycholoGical support After covid-19 InfectioN’ (REGAIN): a structured summary of a study protocol for a randomised controlled trial. Trials 2021; 22: 1–3.

98 Pilloni G, Bikson M, Badran BW, George MS, Kautz SA, Okano AH et al. Update on the Use of Transcranial Electrical Brain Stimulation to Manage Acute and Chronic COVID-19 Symptoms. Front Hum Neurosci 2020; 14: 493.

99 Silva-Filho E, Moura S, da Cruz Santos A, do Socorro Brasileiro-Santos M, de Albuquerque JA. Transcranial direct current stimulation as a strategy to manage COVID-19 pain and fatigue. Rev Assoc Med Bras 2021; 67: 26–28.

100 Gómez L, Vidal B, Cabrera Y, Hernández L, Rondón Y. Successful Treatment of Post-COVID Symptoms With Transcranial Direct Current Stimulation. Prim care companion CNS Disord 2021; 23. doi:10.4088/PCC.21CR03059.

101 Transcranial Direct Stimulation for Persistent Fatigue Treatment Post-COVID-19. 2021.https://www.clinicaltrials.gov/ct2/show/NCT05252481 (accessed 7 Jun2022).

102 Home-based Brain Stimulation Treatment for Post-acute Sequelae of COVID-19 (PASC). 2021.https://clinicaltrials.gov/ct2/show/NCT05092516 (accessed 7 Jun2022).

103 National Institute for Health and Care Excellence. NICE ME/CFS guideline outlines steps for better diagnosis and management. 2021.https://www.nice.org.uk/news/article/nice-me-cfs-guideline-outlines-steps-for-better-diagnosis-and-management (accessed 7 Jun2022).

104 Hughes SE, Haroon S, Subramanian A, McMullan C, Aiyegbusi OL, Turner GM et al. Development and validation of the symptom burden questionnaire for long covid (SBQ-LC): Rasch analysis. BMJ 2022; 377: e070230.

