## Supplementary Material for "Non-pharmacological therapies for post-viral syndromes, including Long COVID: A systematic review"

### Supplementary table 1 – PRISMA checklist

| **Section and Topic** | **Item #** | **Checklist item** | **Location where item is reported** |
| --- | --- | --- | --- |
| **TITLE** | | |  |
| Title | 1 | Identify the report as a systematic review. | 1 |
| **ABSTRACT** | | |  |
| Abstract | 2 | See the PRISMA 2020 for Abstracts checklist. | 3 |
| **INTRODUCTION** | | |  |
| Rationale | 3 | Describe the rationale for the review in the context of existing knowledge. | 5 |
| Objectives | 4 | Provide an explicit statement of the objective(s) or question(s) the review addresses. | 5 |
| **METHODS** | | |  |
| Eligibility criteria | 5 | Specify the inclusion and exclusion criteria for the review and how studies were grouped for the syntheses. | 6 |
| Information sources | 6 | Specify all databases, registers, websites, organisations, reference lists and other sources searched or consulted to identify studies. Specify the date when each source was last searched or consulted. | 6 |
| Search strategy | 7 | Present the full search strategies for all databases, registers and websites, including any filters and limits used. | 6 |
| Selection process | 8 | Specify the methods used to decide whether a study met the inclusion criteria of the review, including how many reviewers screened each record and each report retrieved, whether they worked independently, and if applicable, details of automation tools used in the process. | 7 |
| Data collection process | 9 | Specify the methods used to collect data from reports, including how many reviewers collected data from each report, whether they worked independently, any processes for obtaining or confirming data from study investigators, and if applicable, details of automation tools used in the process. | 7 |
| Data items | 10a | List and define all outcomes for which data were sought. Specify whether all results that were compatible with each outcome domain in each study were sought (e.g. for all measures, time points, analyses), and if not, the methods used to decide which results to collect. | 7 |
|  | 10b | List and define all other variables for which data were sought (e.g. participant and intervention characteristics, funding sources). Describe any assumptions made about any missing or unclear information. | 7 |
| Study risk of bias assessment | 11 | Specify the methods used to assess risk of bias in the included studies, including details of the tool(s) used, how many reviewers assessed each study and whether they worked independently, and if applicable, details of automation tools used in the process. | 7 |
| Effect measures | 12 | Specify for each outcome the effect measure(s) (e.g. risk ratio, mean difference) used in the synthesis or presentation of results. | 7 |
| Synthesis methods | 13a | Describe the processes used to decide which studies were eligible for each synthesis (e.g. tabulating the study intervention characteristics and comparing against the planned groups for each synthesis (item #5)). | 7 |
|  | 13b | Describe any methods required to prepare the data for presentation or synthesis, such as handling of missing summary statistics, or data conversions. | 7 |
|  | 13c | Describe any methods used to tabulate or visually display results of individual studies and syntheses. | 7 |
|  | 13d | Describe any methods used to synthesize results and provide a rationale for the choice(s). If meta-analysis was performed, describe the model(s), method(s) to identify the presence and extent of statistical heterogeneity, and software package(s) used. | 7 |
|  | 13e | Describe any methods used to explore possible causes of heterogeneity among study results (e.g. subgroup analysis, meta-regression). | 7 |
|  | 13f | Describe any sensitivity analyses conducted to assess robustness of the synthesized results. | 7 |
| Reporting bias assessment | 14 | Describe any methods used to assess risk of bias due to missing results in a synthesis (arising from reporting biases). | 7 |
| Certainty assessment | 15 | Describe any methods used to assess certainty (or confidence) in the body of evidence for an outcome. | 7 |
| **RESULTS** | | |  |
| Study selection | 16a | Describe the results of the search and selection process, from the number of records identified in the search to the number of studies included in the review, ideally using a flow diagram. | 9 |
|  | 16b | Cite studies that might appear to meet the inclusion criteria, but which were excluded, and explain why they were excluded. | 8 |
| Study characteristics | 17 | Cite each included study and present its characteristics. | 9-12 |
| Risk of bias in studies | 18 | Present assessments of risk of bias for each included study. | 13 |
| Results of individual studies | 19 | For all outcomes, present, for each study: (a) summary statistics for each group (where appropriate) and (b) an effect estimate and its precision (e.g. confidence/credible interval), ideally using structured tables or plots. | 14-16 |
| Results of syntheses | 20a | For each synthesis, briefly summarise the characteristics and risk of bias among contributing studies. | 10-16 |
|  | 20b | Present results of all statistical syntheses conducted. If meta-analysis was done, present for each the summary estimate and its precision (e.g. confidence/credible interval) and measures of statistical heterogeneity. If comparing groups, describe the direction of the effect. | No statistics undertaken |
|  | 20c | Present results of all investigations of possible causes of heterogeneity among study results. | No statistics undertaken |
|  | 20d | Present results of all sensitivity analyses conducted to assess the robustness of the synthesized results. | No statistics undertaken |
| Reporting biases | 21 | Present assessments of risk of bias due to missing results (arising from reporting biases) for each synthesis assessed. | N/A |
| Certainty of evidence | 22 | Present assessments of certainty (or confidence) in the body of evidence for each outcome assessed. | N/A |
| **DISCUSSION** | | |  |
| Discussion | 23a | Provide a general interpretation of the results in the context of other evidence. | 19 |
|  | 23b | Discuss any limitations of the evidence included in the review. | 19-21 |
|  | 23c | Discuss any limitations of the review processes used. | 21 |
|  | 23d | Discuss implications of the results for practice, policy, and future research. | 19-21 |
| **OTHER INFORMATION** | | |  |
| Registration and protocol | 24a | Provide registration information for the review, including register name and registration number, or state that the review was not registered. | 3 |
|  | 24b | Indicate where the review protocol can be accessed, or state that a protocol was not prepared. | 5 |
|  | 24c | Describe and explain any amendments to information provided at registration or in the protocol. | 6 and 20 |
| Support | 25 | Describe sources of financial or non-financial support for the review, and the role of the funders or sponsors in the review. | 2 |
| Competing interests | 26 | Declare any competing interests of review authors. | 2 |
| Availability of data, code and other materials | 27 | Report which of the following are publicly available and where they can be found: template data collection forms; data extracted from included studies; data used for all analyses; analytic code; any other materials used in the review. | N/A |

### Supplementary figure 1 – Search strategy from MEDLINE

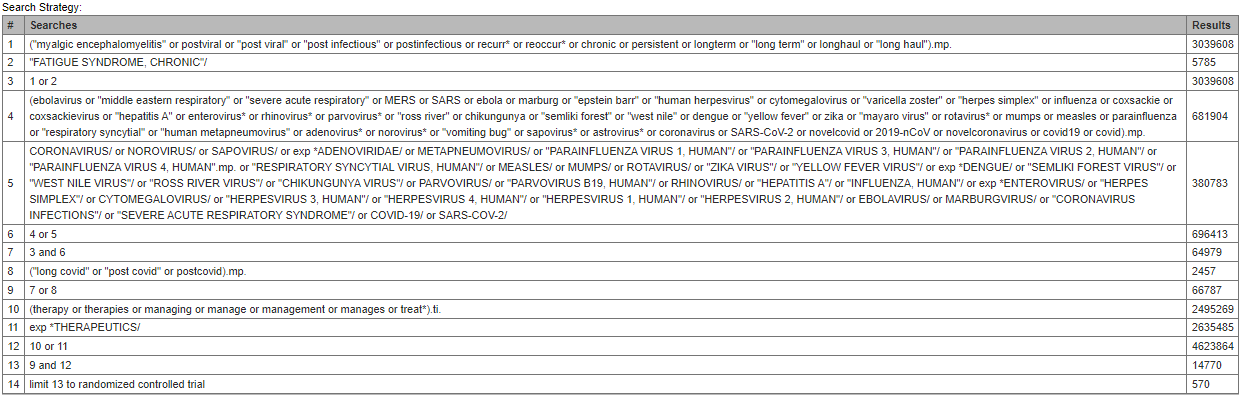

### Supplementary table 2 - GRIPP-2 short form checklist^23^

| Section and topic | Item | Description of involvement |
| --- | --- | --- |
| 1: Aim | Report the aim of PPI in the study | Patients and the public were involved in the development of the research question and design of the review. |
| 2: Methods | Provide a clear description of the methods used for PPI in the study | The PPI activities were supported by patient and public involvement and engagement (PPIE) group and patient partner for the project (JC). We held an initial meeting with members of the PPIE group where members of the research team gave a presentation which highlighted the similarity between post-viral syndromes and Long COVID, the rationale for this review as well as our preliminary ideas. Following the presentation, there was a discussion of the review objectives. The PPIE group members were asked for their thoughts on the review objectives and its scope. Another presentation was given to the PPIE group once the review was completed for their opinions on the findings and recommendations. JC was also involved in the write up of the protocol and final manuscript. |
| 3: Study results | Outcomes—Report the results of PPI in the study, including both positive and negative outcomes | During the design phase the PPIE group confirmed that the objectives were appropriate and comprehensive, they stressed the impact of Long COVID on employment and that the loss of income was very important to patients. JC also provided critical input and opinion during the write up of the protocol independently of PPIE group comments.  Patients and members of the public were not involved in the study selection, extraction, or synthesis phases. |
| 4: Discussion and conclusions | Outcomes—Comment on the extent to which PPI influenced the study overall. Describe positive and negative effects | Therefore, a key goal of this work should be to identify potential non-pharmacological interventions that will facilitate the rehabilitation of patients with long COVID so that they continue to make progress and can adapt to new working methods. Based on their feedback, we ensured that this review reported interventions designed to improve the wellbeing of patients which will facilitate their return to work. |
| 5: Reflections/critical perspective | Comment critically on the study, reflecting on the things that went well and those that did not, so others can learn from this experience | Early engagement with our PPIE group was critical to designing a research question with outcomes relevant to the patient population who will be most affected by any recommendations from this study. |

### Supplementary table 3 – Narrative description for risk of bias assessment

| **de Oliveira et al., 2019** | | |
| --- | --- | --- |
| **Bias** | **Authors’ judgement** | **Support for judgement** |
| Random sequence generation (selection bias) | Low risk | ‘Permuted block randomization was performed using the program Random Allocation 2.0’ |
| Allocation concealment (selection bias) | Low risk | ‘Sealed, opaque, and sequentially numbered enveloped with a 1:1 allocation ratio’ |
| Blinding of participants and personnel (performance bias) | High risk | Blinding of participants was not feasible for this study and therefore there may be risk of placebo effect etc. |
| Blinding of outcome assessment (detection bias) | Low risk | ‘Blinded assessor who did not know to which group each participant had been allocated’ |
| Incomplete outcome data (attrition bias) | Unclear risk | All outcomes were reported on, but intention-to-treat (ITT) analysis was not carried out and it is unknown if this may have biased the results |
| Selective reporting (reporting bias) | Low risk | Outcomes were evaluated against the trial protocol and no discrepancies were found |
| Other bias | Low risk | No other sources of bias were identified |
| **Li et al., 2021** | | |
| **Bias** | **Authors’ judgement** | **Support for judgement** |
| Random sequence generation (selection bias) | Low risk | ‘Permuted allocation sequences for 1:1 block randomization (block size 10-14) stratified by hospital were computer-generated’ |
| Allocation concealment (selection bias) | Low risk | ‘Allocation was concealed by central randomization and only revealed after baseline assessment through call to study center’ |
| Blinding of participants and personnel (performance bias) | High risk | Blinding of participants was not feasible for this study and therefore there may be risk of placebo effect etc. |
| Blinding of outcome assessment (detection bias) | Low risk | ‘Patients and therapists were requested to not disclose allocation to assessors at any time during this study’ |
| Incomplete outcome data (attrition bias) | Low risk | Intention-to-treat analysis was carried out in order to account for those lost to follow up and minimize loss of outcome data |
| Selective reporting (reporting bias) | Low risk | Outcomes were evaluated against the trial protocol and no discrepancies were found |
| Other bias | Low risk | No other sources of bias were identified |
| **Malik et al., 2020** | | |
| **Bias** | **Authors’ judgement** | **Support for judgement** |
| Random sequence generation (selection bias) | Low risk | ‘Participants were randomized to either mental training or care as usual in a 1:1 probability by a computer-based routine for block randomization’ |
| Allocation concealment (selection bias) | Low risk | ‘Allocation concealment was ensured using sequentially numbered, opaque, sealed envelopes’ |
| Blinding of participants and personnel (performance bias) | High risk | Blinding of participants was not feasible for this study and therefore there may be risk of placebo effect etc. |
| Blinding of outcome assessment (detection bias) | Unclear risk | States ‘end-point evaluation was concealed from patients and therapists’, but does not explicitly state whether the outcome assessors were themselves blinded or not |
| Incomplete outcome data (attrition bias) | Low risk | Intention-to-treat analysis was carried out in order to account for those lost to follow up and minimize loss of outcome data |
| Selective reporting (reporting bias) | Low risk | Outcomes were evaluated against the trial protocol and no discrepancies were found |
| Other bias | Low risk | No other sources of bias were identified |
| **Neumann et al., 2021** | | |
| **Bias** | **Authors’ judgement** | **Support for judgement** |
| Random sequence generation (selection bias) | Low risk | ‘One independent examiner performed bloc randomization using randomization.com’ |
| Allocation concealment (selection bias) | Low risk | ‘Results were placed in dark sealed envelopes that were given directly to the examiner responsible for the intervention |
| Blinding of participants and personnel (performance bias) | High risk | Blinding of participants was not feasible for this study and therefore there may be risk of placebo effect etc. |
| Blinding of outcome assessment (detection bias) | Low risk | ‘One independent research assistant tabled and codified the data in order to blind the statistics’ |
| Incomplete outcome data (attrition bias) | Low risk | Intention-to-treat analysis was carried out in order to account for those lost to follow up and minimize loss of outcome data |
| Selective reporting (reporting bias) | Low risk | Outcomes were evaluated against the trial protocol and no discrepancies were found |
| Other bias | Low risk | No other sources of bias were identified |
| **Silva-Filho et al., 2018** | | |
| **Bias** | **Authors’ judgement** | **Support for judgement** |
| Random sequence generation (selection bias) | Low risk | ‘A random numerical sequence was generated ([www.randomization.com](http://www.randomization.com)) to assign each participant’ |
| Allocation concealment (selection bias) | Unclear risk | Unclear if or how allocation concealment was done |
| Blinding of participants and personnel (performance bias) | High risk | ‘Participants and researchers were blind to group allocation throughout the trial’  ‘Sham-tDCS was performed on 5 consecutive days with electrodes placed on the same position’ |
| Blinding of outcome assessment (detection bias) | Low risk | ‘Participants and researchers were blind to group allocation throughout the trial’ |
| Incomplete outcome data (attrition bias) | Low risk | Intention-to-treat analysis was carried out in order to account for those lost to follow up and minimize loss of outcome data |
| Selective reporting (reporting bias) | High risk | Did not report on one of the primary outcomes in the trial protocol (DN4 Questionnaire) |
| Other bias | High risk | ‘The City University of New York has patent on brain simulation with MB as the inventor. MB has equity in Soterix Medical Inc.’ |
